# An observational analysis of patient recruitment in clinical trials in France using real-word database PMSI

**DOI:** 10.1101/2022.04.05.22273463

**Authors:** Joy Seanehia, Aurélie Lampuré, Raimundo Gordejuela, Basma Amdouni, Pedro Manzione, Konstantinos Mammas

**Author notes:** (corresponding author) Joy SEANEHIA. Aurélie LAMPURÉ.

## Abstract

**Background:** France has significant clinical research and development potential, however, struggles in comparison to neighbouring countries. A significant reason is the difficulty to recruit patients, thus causing delays in the availability of new therapies to market. IQVIA uses Health Insurance Claims Data among other data assets, to better locate patients for trials based on the potential of hospitals.

**Objective:** The aim of the study was to monitor whether an increased number of patients enrolled in clinical trials in France was observed when PMSI data supported patient recruitment, as well as describing clinical trial landscape worldwide and in Europe.

**Methods:** We used data from ClinicalTrials.gov and Citeline to describe the clinical trial landscape in Europe between 2010 and 2019. We also looked at the IQVIA internal clinical trial tracker, Clinical Trial Management System (CTMS) to describe IQVIA-run trials and their performance after matching trials supported with PMSI data in France. We compared the average number of enrolled patients per site in PMSI and non-PMSI supported trials according to the study phase, using a Student t-test.

**Results:** Results suggest that the support of PMSI on the average number of enrolled patients per site, when comparing at similar trial phase level, shows a positive trend especially for phase 4 studies (11.0 with PMSI vs 9.3 without PMSI, p=0.67), and for phases 3b, 3 and 1, when compared to non-PMSI supported studies.

**Conclusions:** The findings of this study suggest that PMSI use has the potential to increase patient recruitment into clinical trials run in France, rendering France more attractive in its exploitation of the clinical research potential. Optimising patient recruitment has a direct impact on the availability and timeliness of innovative therapies to market for French patients.

## Introduction

France’s place in the commercial-sponsored clinical trial space has been reducing over a number of years according to the National Doctor’s Academy Working Group, ^1^ despite a high concentration of highly qualified healthcare personnel with strong research capabilities. Some of the reasons for this trend as outlined by the working group, include strict regulations by government bodies, administrative bottlenecks and challenges in recruiting patients for trials in the expected timelines. The working group provided a range of recommendations, one of which is to reduce the burden and costs inefficiencies in patient recruitment.

Improving these efficiencies can be achieved using data-driven patient recruitment means instead of the classic CRO approach, of relying mainly on declarative potential by investigators during outreach activities. Analyses of sites’ enrolment performance vs estimates received from sites during feasibility outreach show that in feasibility assessments, most investigators overestimate their enrollment potential. Based on these overestimations, sites struggle to meet their enrolment targets upon activation, with some remaining non-enrollers throughout the course of the study. Sites struggling with recruitment delays or difficulty finding subjects impacts the power of studies in their ability to demonstrate significant effects. ^2^ Studies taking longer to run or getting discontinued, ^3^ are not only a source of cost and time inefficiencies, but consequently impact the public’s access to innovative therapies.

Even though, several hurdles still impede the patient recruitment process, there has been significant innovation in improving patient recruitment. Targeting these efficiencies in scattered data sources is the platform developed by Mudarankathan et al, ^4^ the Curated Cancer Clinical Outcomes Databases (C3OD) used to centralise the various data and facilitate its access. This platform is intended to speed patient recruitment in oncology studies. Other recent efforts have succeeded in improving efficiencies in patient recruitment without limiting it to therapeutic area. One of such hospital specific information systems is the DARWIN Cohort Management System (DCMS) which identifies patients for genotype matched studies as described by Eubank et al. ^5^ Achieving efficiencies in patient identification and enrolment are underpinned by data and studies like Ni et al, ^6^ explore this potential by using natural language processing and machine learning techniques to match patients to trials. These patient selection approaches contribute to recruitment acceleration and enable rapid generation of valuable scientific research and outcomes to meet public health needs.

However, these approaches are limiting when applied to hospitals that lack the patient potential. Opening sites with no patient potential remain costly and inefficient. IQVIA addresses this gap with a data driven approach in site selection. This task undertaken by a global team relies on local data sources, in-house technology, country-specific expertise, and robust analytics to provide insights on how best to meet this need and ensure that these often-multi-country trials are executed efficiently at the local level.

One of such local data sources used in IQVIA is Health Insurance Claims Data. Similar to all industrialised countries who possess structured payment systems for healthcare costs management, ^7^ France developed *PMSI*, the acronym for Programme de Medicalisation des Systèmes d’Information in 1996, ^8^ following reforms to Hospital Reimbursement and Claims systems. This replacement is based on a coding system known as Pricing by Activity (*Tarification A l’Activité* [T2A]). ^9^ This hospital information system, like many others, is based on Diagnosis Related Groups (DRG) which categorises patients based on their pathologies (*Groupe Homogene de* Malades, [GHM]), representing the average cost of managing these kinds of patients according to their diagnosis, comorbidities and treatment procedures during their hospital stay (*Groupe Homogène de Séjour, [GHS])*. ^10^ This system allows hospitals to receive payment corresponding to their activity and healthcare services delivered. As the primary goal of the data collected with the PMSI system is to ensure hospital payments, patient data is anonymised and centralised at the national level, with the governing body in charge of hospital data, *Agence Technique de l’Information sur l’Hospitalisation* (ATIH) overseeing the process. ^11^

The current exploitation of this data goes beyond its primary purpose of hospital budget allocation, nevertheless, its use is seen in several medical research fields, and Boudemaghe et al. ^8^ outline a list of such research areas, which span various disease indications. ^12, 13, 14^

In 2019 the ATIH granted the R&D Business Unit within IQVIA, access to PMSI. The rigor that accompanies the authorisation to this database is ensured via a double authentication system, which guarantees patient data protection and governance. This process is managed by the body, Centre d’accès sécurisé aux données (CASD). ^15^ This database’s coverage is extensive, and records patients’ characteristics and comorbidities. The following information is available in PMSI database: 1) hospital data (entity identifier (Finess), geographical/legal entity identifier), 2) patient administrative information (age, gender, patient identifier), 3) patient medical information (primary diagnosis, related diagnosis and associated significant diagnosis (ICD10 codes), procedures, medical devices and drugs (“liste-en-sus”)), and 4) hospitalization information (DRG, length of hospitalisation). This record tracks hospital admissions in various units like the Medicine, Surgery and Obstetric units (French acronym MCO for Médecine, Chirurgie, Obstétrique), Psychiatry and Home Hospitalisation (HAD). It covers all hospitalisations in public and private healthcare centres (inpatient overnight stays, day hospitalisations and outpatient consultations). To further prevent the identification of patients during Site Identification for Patient Density, their aggregation at the site/hospital level is carried out in a secure environment.

IQVIA’s access to the French Claims dataset as an addition to the panel of data capabilities, one of the 4 pillars of IQVIA Core, distinguishes it in its execution of clinical research and development. A key advantage of PMSI in limiting selection bias is its coverage of private, public and private-with-public-interest (établissement privé d’intérêt collectif, ESPIC) sector hospitals.

The above-mentioned strengths of the French Claims data, as well as its ability to bridge the gap in the French clinical trial space, could help France become more attractive for industry-sponsored trials.

In this study, we monitored whether an increased number of patients enrolled in clinical trials in France was observed when PMSI data supported patient recruitment, as well as describing clinical trial landscape worldwide and in Europe. First, the objective was to describe patient enrolment in IQVIA clinical trials, worldwide over the period 2010-2019, and focus into Europe, to compare contribution of the main European countries in the industry-sponsored clinical trial space (the latter accounting for 50% of trials in France). ^16^ Secondly, the objective was to monitor in France, the average number of patients enrolled by site in trials supported by PMSI data vs those not supported by PMSI data, according to the trial phase.

## Methods

We investigated the European clinical trials activity using two large clinical trial databases, ClinicalTrials.gov, ^17^ and Trialtrove from Citeline, ^18^ which are databases of privately and publicly funded clinical studies conducted around the world. We also used IQVIA’s internal clinical trial tracker, Clinical Trial Management System (CTMS) to locate hospitals/sites and to measure their performance indicators. CTMS as exists in various Contract Research Organisations (CROs), contains data on past and ongoing trials and is updated with hundreds of data points at the site level. Python version 3.7.9, R version 3.6.1, and SAS version 9.4 were used in the development environment. To investigate the industry-sponsored clinical trials in Europe, we submitted queries to ClinicalTrials.gov and Citeline via their website, allowing for the extraction of datasets from these databases, in June 2021 (authorisations were provided for both databases). On ClinicalTrials.gov, we specified three criteria for the extraction: the country (list of European countries selected available in Supplementary Table 1), the funder type “Industry” and the study start dates “from 01/01/2010 to 12/31/2019”. On the Citeline website, on the Trialtrove page, the same three criteria were specified. Both databases were pooled, and duplicates removed based on NCT number and title. A graph with the percentage of trials operated in the top 5 main European countries among all industry studies run in Europe was generated and presented.

Overall patient enrolment at IQVIA sites across the world was plotted using data from our CTMS database. Three sub-regions were highlighted: The United States of America, NEMEA (Northern Europe, Middle East and Africa) and CESE (Central and Eastern Europe) with the latter two being part of the EMEA (Europe, Middle East and Africa) region. We considered clinical trials with sites enrolled between January 2010 and December 2019 in the analysis, including studies from all four clinical trial phases. Extensive data cleaning preceded this step consisting of the removal of outliers. We liaised with the clinical team on considerations of outliers where unclear from a statistical perspective. Their input guided decisions on such outliers, as no basic statistical methods sufficed to deal with them, given their heterogeneity based on trial phase, therapeutic area and size of the hospital.

The enrolment trend in the EMEA region was then displayed for the highest-ranking countries by patient number, also covering a 10-year span from 2010-2019. The choice of variable as a time indicator, useful for measuring the start of clinical activity at site level was the “*Activation Date*”.

To test and validate the assumption that patient potential at hospitals based on their density, positively impact clinical research and development in France, the studies were then matched with those supported with PMSI data, following an extraction from an internal data repository of all studies supported by the R&D Business Unit.

We performed descriptive analysis and hypothesis tests to monitor whether an increased number of subjects enrolled per site is observed in studies with PMSI support versus those without. Student’s t-test was used to compare the means of patients enrolled per site, in studies supported by PMSI data vs those without, stratified by phase due to different methodologies involved.

## Results

### Clinical Trial Landscape

Data extracted from CTMS covered the period ranging from January 2009 till January 2021. During this period, 13,402 clinical trials at 1,186,396 sites were tracked in CTMS database. The average number of subjects enrolled per site was 3.62 (SD=32.44).

Fig 1 displays IQVIA patient enrolment at sites across the world categorised by regions and sub regions. The United States of America, the NEMEA and CESE subregions have the highest representation in terms of patients enrolled and studies run. The US records 1,600 studies with almost 250,000 patients during the period 2010-2019 in IQVIA run trials.

**Fig 1.**
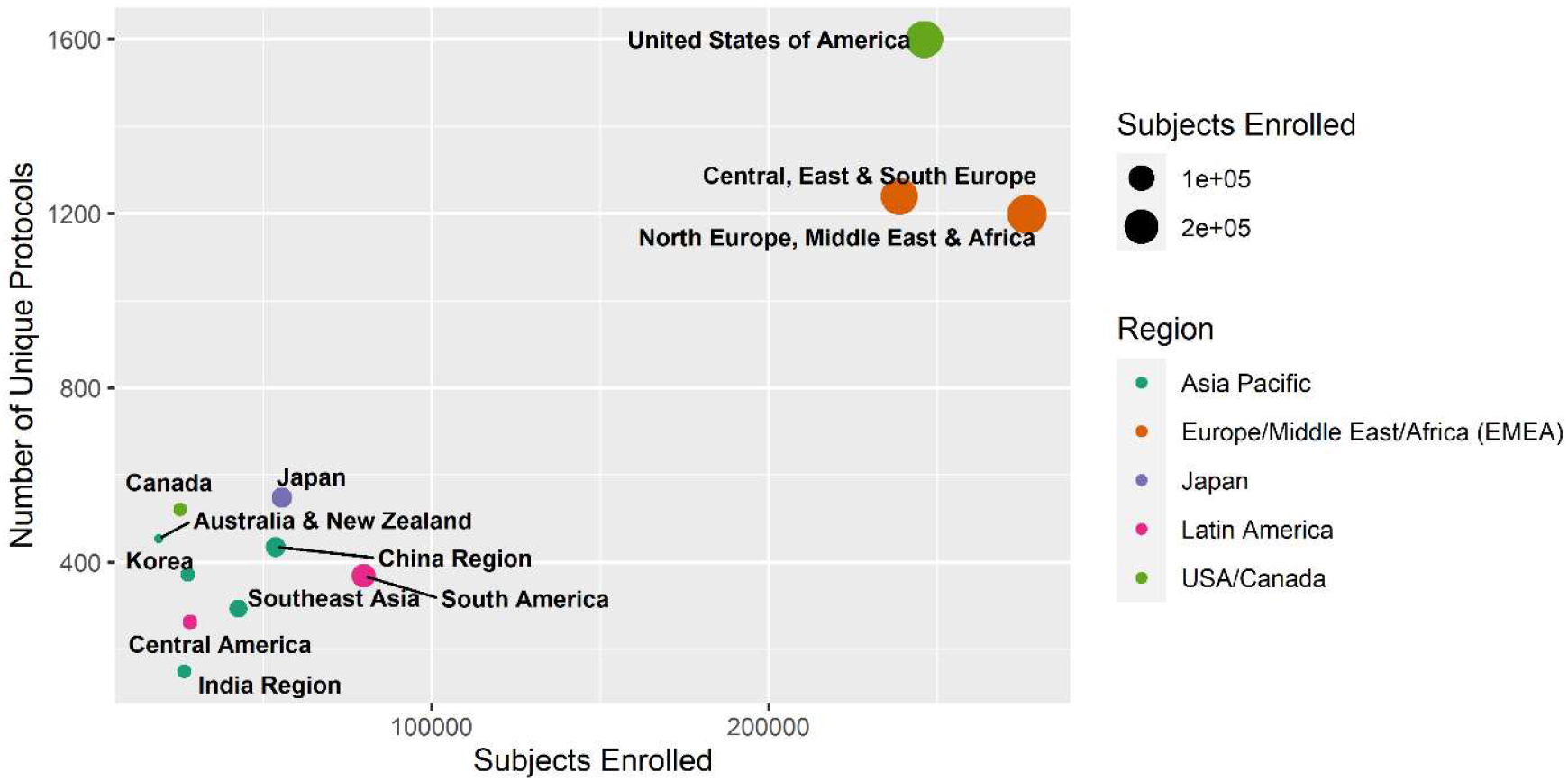
Patient enrolment in IQVIA clinical trials worldwide 2010-2019, CTMS, IQVIA France.

The share of 5 European countries and their contributions to the industry-sponsored clinical trial activity is detailed in Fig 2, over the course of ten years. This figure illustrates the proportion of each country’s participation in clinical trials, in comparison to their counterparts in the European region, regardless of other region’s participation in said studies. This is based on Citeline and CT.gov databases. The contribution of each country is calculated by dividing the number of clinical trials a country had participated in by the total number of clinical trials that took place in Europe (with or without other regions involved). Germany appears as the main contributor since 2010, with a decreasing trend over the years (41% in 2010; 35% in 2019), closely followed by the United Kingdom (UK) from 2016-2019. France is in third place, contributing on average 27%-31% of clinical trials in the past decade, close to Spain which has been continuously increasing over the course of the past few years. Italy has been participating in almost one fourth of clinical trials in Europe for several years.

**Fig 2.**
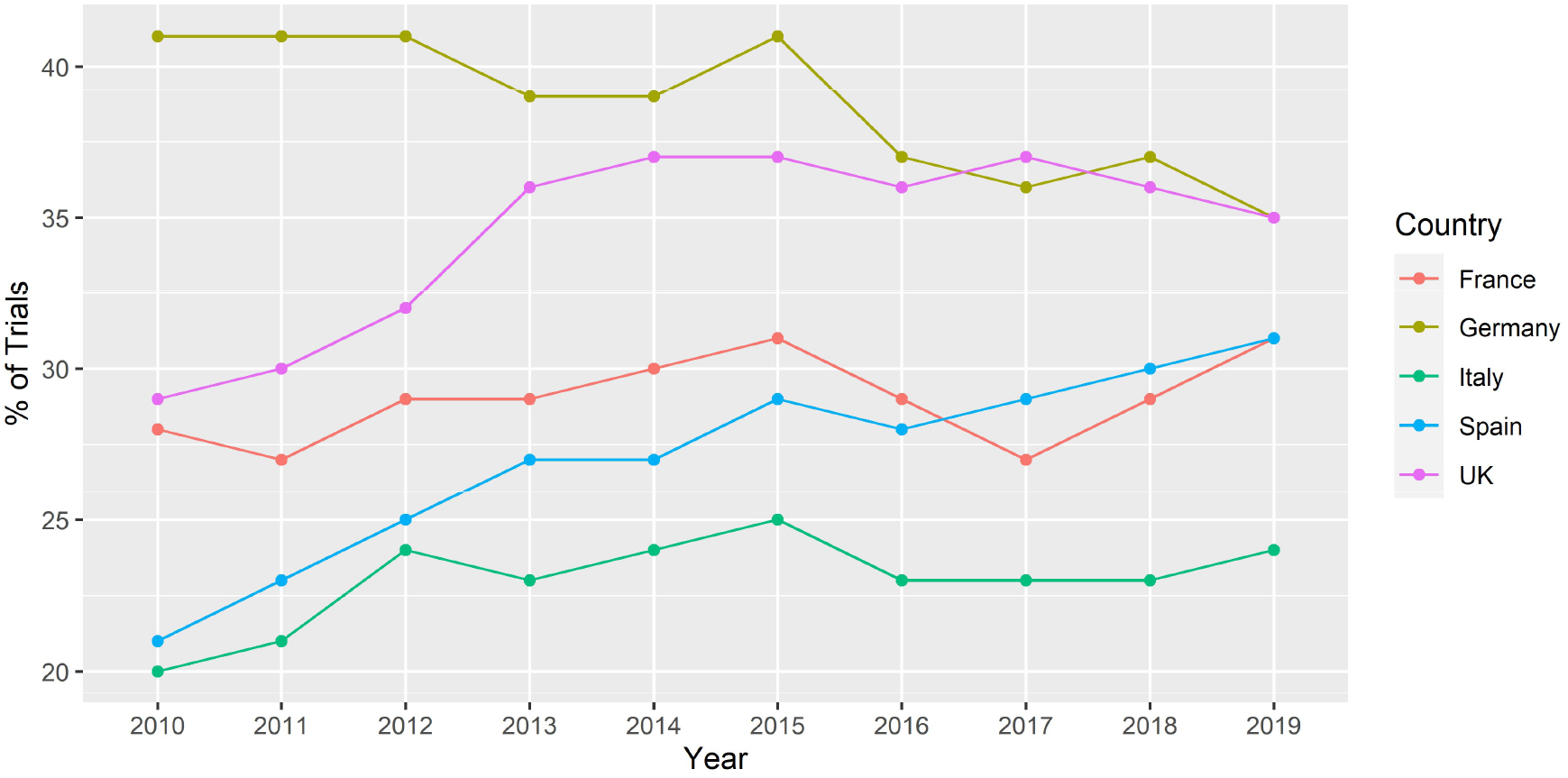
Percentages of main contributing European countries in the industry-sponsored clinical trial landscape, 2010-2019, Citeline and CT.gov.

CTMS seems to be consistent with CT.gov and Citeline databases, as they highlight that France is in a dynamic region with neighbouring countries (UK, Germany and Italy) ranking in the top 5 countries in patient recruitment from 2010-2019. As seen in Fig 3, the UK has 46,638 patients enrolled during that period in IQVIA-run trials and appears as the leader in Europe.

**Fig 3.**
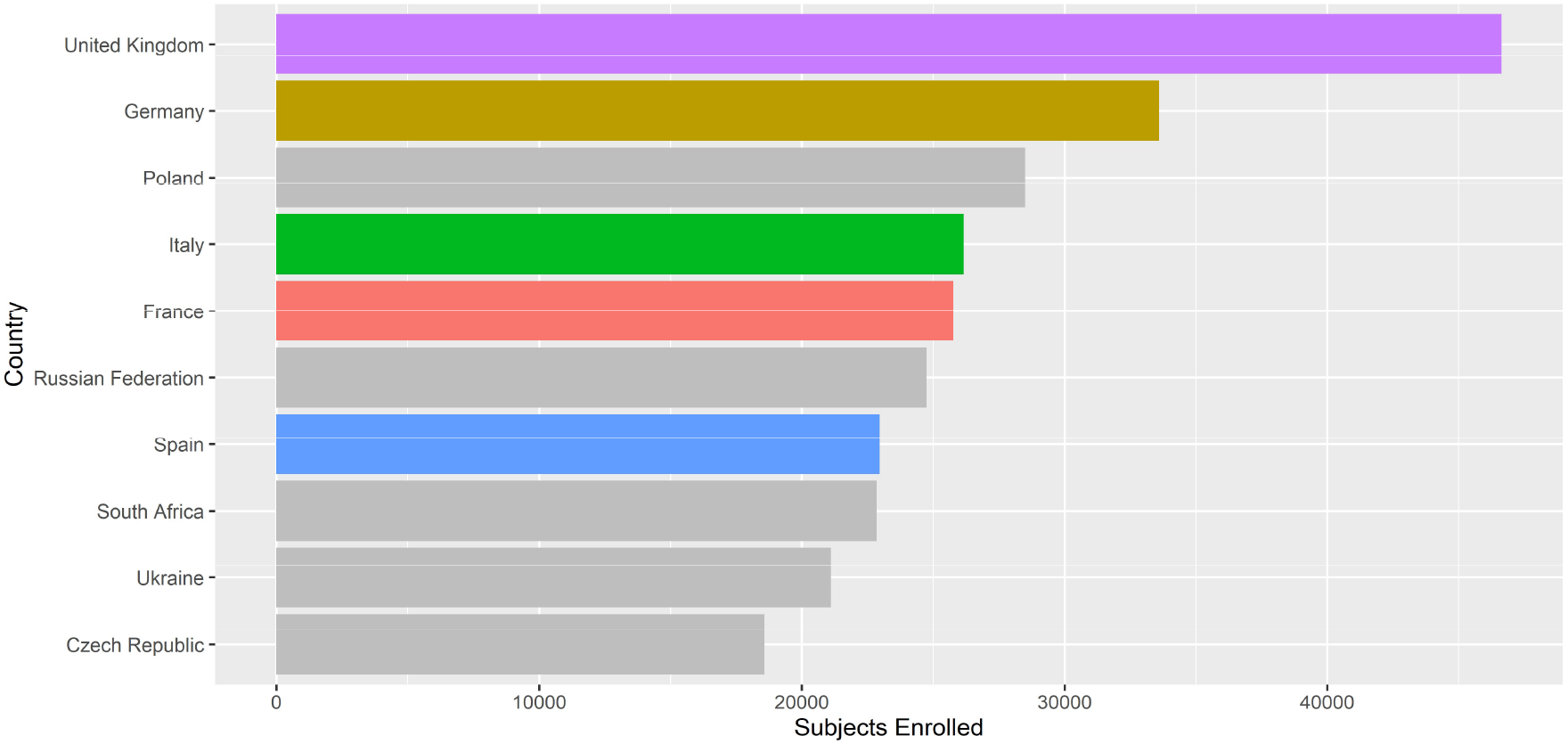
Patient enrolment in EMEA by country, 2010-2019, CTMS, IQVIA France.

### Patient Recruitment on Trials supported with PMSI data

To understand whether an increased number of patients enrolled is observed in trials supported with PMSI data, this analysis focused on studies and sites, both present in CTMS and PMSI databases, accounting for 47% of the available records in CTMS. We considered this as the reference database as it is used to derive the experiments described in this study. In addition, we performed quality control on the available data and records with missing values were excluded from the analysis.

IQVIA’s activity covered 10% of the French clinical trial market over the past 10 years, after analysis of the trial share of CT.gov and Citeline recorded trials attributed to IQVIA. Among the 2,879 sites initiated for IQVIA studies in France between 2010 and 2019 and reported in CTMS, 47% of sites had the number of subjects screened collected, and 26% of sites had the site activation date recorded in the database.

Among the 135 IQVIA-run studies from 2017-2018 that included French sites and have available data in CTMS (activation dates and number of enrolled patients), 29 of them were supported with PMSI data. A comparison of the average number of enrolled subjects by phase is plotted in Fig 4.

**Fig 4.**
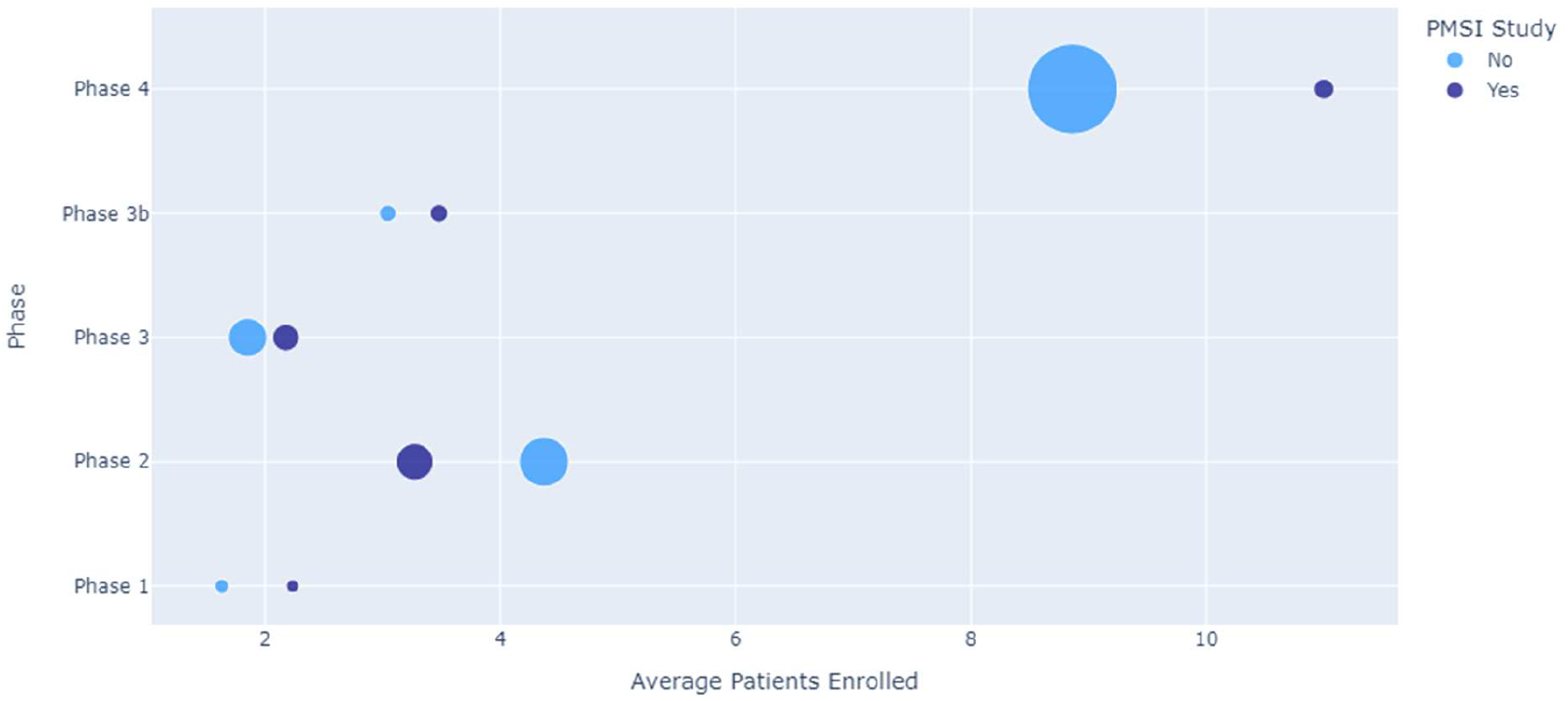
**Average Patients Enrolled in French sites supported or not by PMSI data by study phase, 2017-2018, CTMS, IQVIA France (size of circle is proportional to the absolute number of patients included in each phase)**

A comparison of means performed by phases does not demonstrate statistical significance, but a positive trend is seen among studies supported with PMSI data at sites activated in 2017-2018, especially in phase 4 clinical trials, with on average two more patients recruited in PMSI supported studies (11.0 with PMSI vs. 9.3 without PMSI, p=0.67). A slight positive trend is also present in phases 3 (2.2 with PMSI vs. 1.9 without PMSI, p=0.42), 3b (3.5 with PMSI vs. 3.0 without PMSI, p=0.69) and 1 (2.2 with PMSI vs. 1.6 without PMSI, p=0.47). Phase 2 clinical trials did not provide the same result with PMSI data use (3.3 with PMSI vs. 4.4 without PMSI, p=0.41). Table 1 details the study sample size of those supported by PMSI vs those not supported by PMSI. These studies are stratified by trial phase.

**Table 1.**
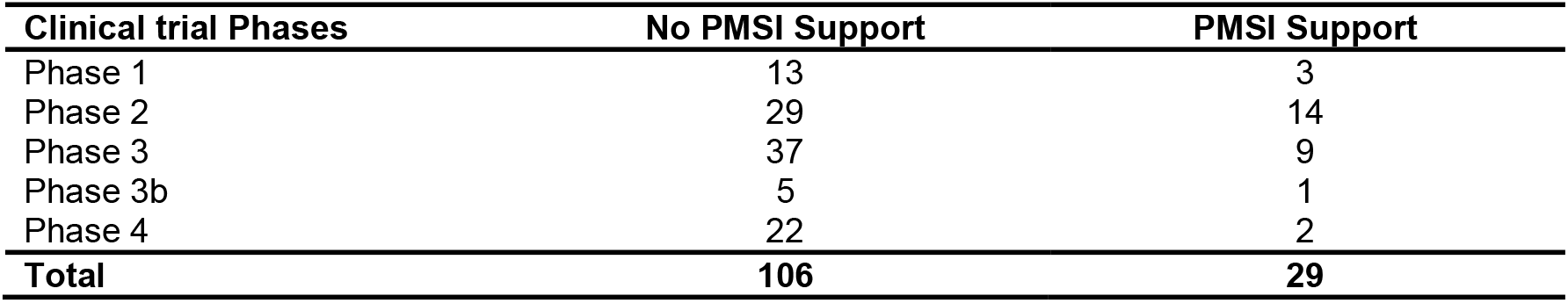
Number of studies included for each phase.

| **Clinical trial Phases** | **No PMSI Support** | **PMSI Support** |
| --- | --- | --- |
| Phase 1 | 13 | 3 |
| Phase 2 | 29 | 14 |
| Phase 3 | 37 | 9 |
| Phase 3b | 5 | 1 |
| Phase 4 | 22 | 2 |
| **Total** | **106** | **29** |

## Discussion

In this study, we monitored the number of patients enrolled in trials, with PMSI and without PMSI data support, during the period 2017-2018. Although the results of the hypotheses tests suggested that the number of patients enrolled to clinical trials with PMSI data support did not increase significantly, a slight increase of patient recruitment was observed during the reference period. PMSI is already used extensively at IQVIA in various medical research fields, and it is usually employed to strategically select sites and accelerate patient recruitment, in order to get innovative therapies to patients in a timely manner. IQVIA’s access to this database for clinical trial purposes has been allowed since 2019, and this publication investigates the patient recruitment trend of this data till date.

IQVIA’s activity was particularly explored in France for the period during which IQVIA supported clinical trial study teams with analytics based on the patient potential, to identify sites with recruitment potential derived from PMSI data. PMSI data was matched to data from the IQVIA trial tracker, CTMS, and a positive trend was observed between the enrolment of patients in PMSI supported trials in clinical phases 1, 3, 3b and 4. Several hypotheses could explain this trend.

The number of patients enrolled in clinical trials supported with PMSI data was investigated when the analysis was limited to studies supported in 2017 and 2018, and when analyses were carried out by phase. The timeline was limited to 2018 as studies supported after 2018 were mostly meant to recruit in 2020, a year which saw most clinical activity reduced or halted due to Covid-related-activity. The results seen from IQVIA’s access to the PMSI data and its positive association with recruitment especially in phase 1,3, 3b and 4 could be useful in drafting recommendations to health authorities to guide their policy decision-making in a data-driven manner. This potential will help maintain access to this data which brings innovative therapies to French patients in a more efficient and timely manner. This paper also explored the clinical trial activity during 2010-2019 and identified the top contributing countries in the EMEA region. IQVIA’s clinical trial landscape was compared to the general activity of clinical trials based on various geographies and timelines, relying on data for the latter derived from Citeline and CT.gov. Germany and the UK appeared as leaders in terms of number of clinical trials and number of patients recruited. The UK was the leader in IQVIA-run trials, with the highest number of enrolled patients in Europe. A recent report from the Association of the British Pharmaceutical Industry (ABPI) confirms that the UK is a world-leader in clinical research for heart disease, immunology and conditions affecting the nervous system. ^19^

## Limitations

PMSI provides information about patient volume, but this is only one of the parameters considered for site identification and site selection. There are additional important factors (e.g. clinical research experience, quality scores, etc) taken into consideration in the site selection process other than patient volume, which could dilute the impact that PMSI data has on the final site list. In this study we monitor 10 years of the clinical trial landscape, but we have only two and a half years of PMSI data availability. Finally, PMSI analytics only supports a fraction of IQVIA-awarded studies at that period.

In addition, the low sample size and the lack of statistical power in the analyses does not allow us to perform an analysis at therapeutic area level. It could be interesting in a few years, with larger time coverage and sample size, to perform this additional analysis. Indeed, we could then assume that depending on the indication, PMSI support might be more necessary for some studies than other.

CT.gov and Citeline databases enabled the investigation of the global trend in the clinical trial landscape. However, CT.gov and Citeline databases only provide information at the study level, unlike CTMS, they did not allow any comparison at the more granular site level regarding site activation dates, number of subjects enrolled by site, etc, which was the objective in this article. These two major databases have good coverage of past and ongoing clinical studies, whereas CTMS data is missing considerable data at certain data points possibly because these are secondary to the activity of the clinical team.

## Conclusion

As clinical trials are the main drivers in advancing innovation in patient care and treatment pathway, optimising patient recruitment has a direct impact on the availability and timeliness of innovative therapies to market. PMSI data allows this possibility, though the evidence so far shows this positive trend in phases 1,3, 3b and 4 studies. This knowledge on the phase-specific impacts could help in focusing on these specific trials, while looking at other solutions of patient recruitment improvement for the remaining phase.

## Data Availability

All data produced in the present study are available upon reasonable request to the authors

## Acknowledgements

We are grateful to our colleagues for their helpful feedback and perspectives on the paper especially Johanna Van Caneghem, Geoffray Bizouard for his PMSI expertise and Cyrille Leroy. The author RG has changed affiliations since his contributions to the paper. He now works in the EMEA Commercial Effectiveness Services, Consulting Services in Madrid, Spain.

## References

1. Lebranchu Y BMCea. La place de la France dans les essais cliniques à promotion industrielle. Bulletin de l’Académie Nationale de Médecine. 2018;202(5):837–857.

2. Swanson GM WA. Recruiting minorities into clinical trials: toward a participant-friendly system. J Natl Cancer Inst. 1995 Dec 6;87(23).

3. Walson PD. Patient recruitment: US perspective. American Academy of Pediatrics. 1999;104:619–22.

4. Mudaranthakam DP TJHJPDCSPMFBGBKDMM. A Curated Cancer Clinical Outcomes Database (C3OD) for accelerating patient recruitment in cancer clinical trials. JAMIA Open. 2018 Oct;1(2):166–171.

5. Eubank MH, Hyman DM, Kanakamedala AD, Gardos SM, Wills JM, Stetson PD. Automated eligibility screening and monitoring for genotype-driven precision oncology trials. J Am Med Inform Assoc. 2016 Jul;23(4).

6. Ni Y, Kennebeck S, Dexheimer JW, et al. Automated clinical trial eligibility prescreening: increasing the efficiency of patient identification for clinical trials in the emergency department. Journal of the American Medical Informatics Association. January 2015;22(1):166–178.

7. Landais P, Boudemaghe T, Suehs C, Dedet G, Lebihan-Benjamin C. Computerized medico-economic decision making: an international comparison. Paris: Springer Paris; 2014.

8. Boudemaghe T, Belhadj I. Data Resource Profile: The French National Uniform Hospital Discharge Data Set Database (PMSI). International Journal of Epidemiology. April 2017;Volume 46(2):392–392d.

9. Ministère des solidarités et de la santé. Financement des établissements de santé. Ministère des Solidarités et de la Santé. July 2, 2021. Available at: https://solidarites-sante.gouv.fr/professionnels/gerer-un-etablissement-de-sante-medico-social/financement/financement-des-etablissements-de-sante-10795/article/financement-des-etablissements-de-sante.

10. ATIH. Manuel des groupes homogènes de malades. ATIH. March 1, 2021. https://www.atih.sante.fr/sites/default/files/public/content/4018/volume_1.pdf.

11. ATIH. missions. https://www.atih.sante.fr/l-atih/. July 9, 2021. Available at: https://www.atih.sante.fr/l-atih/missions.

12. Maura G BPBKBCRPAFZM. Comparison of the short-term risk of bleeding and arterial thromboembolic events in nonvalvular atrial fibrillation patients newly treated with dabigatran or rivaroxaban versus vitamin K antagonists: a French nationwide propensity-matched cohort study. Circulation. SEPTEMBER 2015;132(13):1252–60.

13. Piroth L, Cottenet J, Mariet AS, et al. Comparison of the characteristics, morbidity, and mortality of COVID-19 and seasonal influenza: a nationwide, population-based retrospective cohort study. The Lancer Respiratory Medicine. 2021;9(3):251–259.

14. Sparsa A, Loustaud-Ratti V, Mousset-Hovaere M, et al. Syndrome d’hypersensibilité médicamenteuse en pratique interniste : pièges diagnostique et thérapeutique.Huit observations. La Revue de Médecine Interne. 2000;21(12):1052–1059.

15. CASD. CASD. Les sources de données déjà disponibles au CASD. July 2, 2021. Available at: https://www.casd.eu/les-sources-de-donnees-disponibles-au-casd/.

16. Atal I, Trinquart L, Porcher R, Ravaud P. Differential Globalization of Industry-and Non-Industry–Sponsored Clinical Trials. Plos One. 2015;10(12):e0145122.

17. ClinicalTrials.gov. ClinicalTrials.gov. July 2, 2021. Available at: https://www.clinicaltrials.gov/.

18. Citeline. Trialstrove. July 2, 2021. Available at: https://citeline.informa.com/trials/results.

19. ABPI. Clinical trials: How the UK is researching medicines of the future. 2019. https://www.abpi.org.uk/publications/clinical-trials-how-the-uk-is-researching-medicines-of-the-future/.

