## Supplementary Table 1. for "An observational analysis of patient recruitment in clinical trials in France using real-word database PMSI"

### Supplementary Materials

**Supplementary Table 1. List of European countries considered in CT.gov and Citeline requests**

| European countries | | |
| --- | --- | --- |
| Albania | Georgia | Montenegro |
| Andorra | Germany | Netherlands |
| Armenia | Greece | Norway |
| Austria | Hungary | Poland |
| Azerbaijan | Iceland | Portugal |
| Belarus | Ireland | Romania |
| Belgium | Italy | San Marino |
| Bosnia and Herzegovina | Kosovo | Serbia |
| Bulgaria | Latvia | Slovakia |
| Croatia | Liechtenstein | Slovenia |
| Cyprus | Lithuania | Spain |
| Czech Republic | Luxembourg | Sweden |
| Denmark | Macedonia | Switzerland |
| Estonia | Malta | Ukraine |
| Finland | Moldova | United Kingdom |
| France | Monaco | Vatican City |
